# Do patients comply with 12-hour lithium blood level timing? Findings from a controlled clinical trial and a real-world clinical setting

**DOI:** 10.1101/2024.10.17.24315010

**Authors:** Søren L. Jacobsen, Christian L. Kraft, Andrew A. Nierenberg, Torben A. Devantier, Ole Köhler-Forsberg

**Author notes:** Corresponding author: Psychosis Research Unit; Aarhus University Hospital Psychiatry; Palle Juul-Jensens Boulevard 175; 8200 Aarhus; Mail.

## Abstract

**Background:** Lithium blood levels are measured routinely 12 hours after lithium dosing, but no study has evaluated the actual compliance of patients with the 12-hour levels.

**Methods:** First, we used the US multicenter clinical trial “Bipolar CHOICE” (n=145 patients treated with lithium with N=287 lithium blood tests), where participants reported the time since the last lithium dose at lithium blood tests. Second, we included all lithium blood tests (3,179 individuals, 52,837 blood tests) from hospitals and private practitioners in the Central Denmark Region (CDR, approximate population 1.3 million) during 2012-2022 including time of lithium blood tests and the registered time when patients were supposed to take their lithium dose.

**Results:** In Bipolar CHOICE, participants took the lithium blood test at a mean/median of 12.8/12 hours (SD=9.1, IQR=10.5-14) after the lithium dose, but the range was 0.5-120 hours and 44.9% had the blood test taken <10 or >14 hours after the lithium dose. Those with a blood test >14 hours after the lithium dose had significantly lower lithium levels (0.41 vs. 0.64). In the CDR, the mean/median time was 14.5/13.7 (SD=3.8, IQR=12.0-15.8) and 49.7% had the blood test taken <10 or >14 hours after the supposed intake of lithium. Those with >16 hours between lithium intake and the lithium blood test were more often followed by general practitioners and showed higher creatinine concentrations.

**Conclusions:** Approximately half of lithium blood tests do not comply with guideline-based recommendations for 12-hour trough levels, emphasizing the need for solutions solving this clinical need.

## Background

Lithium represents a first-line mood-stabilizing treatment option for bipolar disorder (1-4). Treatment dosages are based on the clinical effect and lithium blood levels (4,5), which can be measured easily and accurately. Guidelines recommend measuring lithium blood levels during dose changes but also regularly when a patient is on a stable dose (4,6,7). Furthermore, lithium levels are measured to avoid high concentrations and hence potential side effects of lithium treatment, such as long-term effects on the kidneys (8-14).

Standard practice for >50 years has been that lithium levels should be measured 12 hours after the patient took the last lithium dose (15), with most guidelines recommending a window between 10 and 14 hours (4,6,7). Standard practice is that the patient should take the lithium dose in the evening and the blood test the next morning before a possible morning dose. This timing is important as the lithium concentration varies considerably in the time between two lithium intakes. However, not only is adherence to lithium treatment low (16) and regular therapeutic drug monitoring often not performed (17,18), but many patients find it difficult to follow the instructions to take their lithium 12 hours before a blood test. Hence, several factors may contribute to a discrepancy between the actual 12-hour lithium level and the measured lithium concentration. Notwithstanding, the latter provides a basis for clinical decision-making.

Despite the clear clinical importance of assaying the 12-hour lithium level, no study has evaluated whether patients actually comply with the recommended 12-hour blood test timing. We aimed to investigate the compliance with standard timing guidelines for lithium blood tests by using a large representative clinical trial and an observational study population of lithium-treated patients.

## Methods

### Setting

This study is based on two representative cohorts of patients with bipolar disorder with distinct advantages, being 1) the clinical “Bipolar CHOICE” US multicenter trial with specific patient-reported information on the time of the last lithium dose, and 2) a Danish observational setting representing data from everyday clinical work in the Central Denmark Region (CDR). The CDR is one of five administrative regions in Denmark responsible for the healthcare system (approximate population 1.3 million).

### Participants in Bipolar CHOICE

The Bipolar CHOICE trial (24) was a 24-week multi-site, randomized comparative effectiveness trial conducted at 11 US-sites, comparing lithium carbonate to quetiapine. Subjects provided verbal and written informed consent prior to participation and all Institutional Review Boards of the different sites approved the study protocol. The rationale, design, and specific methods of Bipolar CHOICE are reported in detail elsewhere (24).

For Bipolar CHOICE, 692 patients aged between 18 and 62 years were screened, whereof 482 were randomized. A total of 240 individuals were randomized to lithium carbonate. Bipolar CHOICE applied broad inclusion and limited exclusion criteria to maximize heterogeneity of the sample and generalizability of the results. Participants were at study entry required to have a DSM-IV-TR bipolar I or bipolar II diagnosis and to be at least mildly symptomatic (Clinical Global Impression Scale for Bipolar Disorder (CGI-BP) ≥3 (25)). The bipolar disorder diagnosis was determined using the extended Mini-International Neuropsychiatric Interview, an electronic version of a validated structured diagnostic interview. Clinical interviews obtained demographic information and medical history.

### Lithium blood levels and time since last lithium dose in Bipolar CHOICE

Among the 240 patients randomized to lithium in the Bipolar CHOICE trial, lithium levels were measured via an antecubital vein blood sample after 2, 16 and 24 weeks of treatment at normal follow-up visits. Lithium levels are expressed in milliequivalents per liter (mEq\L). At these visits, patients reported the number of hours since their last lithium dose before the blood test. Furthermore, levels of creatinine were measured at baseline (i.e., week 0) and after 16 and 24 weeks.

The target lithium concentration was 0.6 mEq\L, corresponding to an approximate dose of 900 mg. Patients started with lithium treatment at study entry; hence, the 2-week visit represents a visit during up-titration. Most patients were on their stable maintenance dose at weeks 16 and 24.

### Participants in the CDR

In the other part of the study, we used data from the CDR, which has registered information on the time point when patients were supposed to take their lithium dose and the time stamp for the lithium blood test. Since 2012, all blood tests in the CDR at hospitals and private practitioners (i.e., including both private practicing psychiatrists and general practitioners) have been registered, thus covering all lithium blood tests in the CDR. Furthermore, information on all prescribed medications is available, such as lithium, but without information on specific doses. Approval for the present study was granted by the Legal Office in the Central Denmark Region in accordance with the Danish Health Care Act §46, Section 2. According to Danish Law, patient consent and approval from ethical/institutional review boards are not required for studies of this type.

We identified all individuals with available data on lithium blood testing and supposed time of lithium intake during the period from January 1, 2012, to July 31, 2022. This resulted in a total of 3,779 patients and 52,837 blood tests measuring lithium and/or creatinine. We excluded some blood tests. First, we only included blood tests taken between 6 am and 6 pm to restrict to routine samples and exclude acute blood tests taken in connection with hospitalization or emergency room contacts (e.g., due to suspected lithium poisoning), which resulted in an exclusion of 446 blood tests. Second, we restricted to blood tests with P-lithium values <1.5 mmol/l to avoid cases of lithium-poisoning, resulting in an exclusion of 321 blood tests.

### Lithium blood levels and time since last lithium dose in the CDR

Among the remaining 52,070 lithium blood tests, we calculated the time between the supposed lithium intake (e.g., at 8 pm) and the time stamp for the blood-test (e.g., at 9.30 am) and used this as a proxy for the time in hours between the lithium intake and the lithium blood test (in this example 13.5 hours, i.e., from 8 pm to 9.30 am). This was necessary as we only had the hourly timestamp for the supposed lithium intake, and not the patient-reported time of lithium intake.

In the case of individuals with a morning dose of lithium, we calculated the time between the most recent dose (e.g., 8 pm the previous evening) and expected that individuals did not take the morning dose before the lithium blood test, as recommended by guidelines.

### Statistical analysis

All analyses were performed using STATA version 18.0. Variables are expressed as percentages and mean or median values including standard deviations (SD) or interquartile ranges (IQR).

First, we calculated the mean and percentages of blood tests that were taken 12 hours after the most recent lithium dose. This was self-reported in Bipolar CHOICE, while in the CDR, this was defined as lithium blood tests taken between 11.5 and 12.5 hours after the supposed lithium intake.

Second, we calculated the number of lithium blood tests that were taken within the guideline-recommended interval of 10 to 14 hours after the last lithium dose.

Third, we plotted the measured lithium blood concentration and the time since the last lithium dose (i.e., patient-reported in Bipolar CHOICE and calculated in the CDR) to illustrate the correlation between these two factors. In the CDR data, we plotted this depending on whether the lithium blood test was ordered from a psychiatric hospital setting (i.e., an inpatient or outpatient psychiatric hospital department) or by non-hospital settings (i.e., private psychiatrists or other private practitioners, with the vast majority being general practitioners). The purpose of this differentiation was to evaluate if the relation between the lithium blood concentration and time since last lithium intake was different in these two settings. A priori, we speculated that there is less awareness about and hence attention to taking lithium blood tests as close to 12-hours after last lithium intake as possible in non-hospital settings than in psychiatric hospital settings. Therefore, clinicians in non-hospital settings would also be less likely to take into account any significant deviations from the 12 hours when interpreting lithium blood test results. Patients in a non-hospital setting would thus be at higher risk of having their lithium blood tests taken considerably later than 12 hours after last lithium intake, with this falsely low lithium level representing the basis for clinical decision-making on the lithium dose. As a consequence, a higher number of patients may be on a higher lithium dose than intended in a non-hospital compared to a psychiatric hospital setting.

Fourth, in the CDR data, we furthermore plotted the time in hours between the supposed lithium intake and the lithium blood test (e.g., 16 hours) depending on the last supposed lithium intake (e.g., 8 or 10 pm). This was done to show patterns of the supposed lithium intake in a real-world setting and to illustrate whether there are differences in the time between lithium intake and lithium blood tests depending on the time of day the patients are supposed to take their lithium (e.g., how many do take lithium at time points that complicate correct 12-hour blood sampling, such as only a morning dose or an evening dose at 6 pm).

Finally, in the CDR, we calculated the mean (SD) time between the supposed last lithium intake and the lithium blood test stamp depending on calendar year (i.e., 2012, 2013, etc.) to check for potential differences over time.

## Results

### Bipolar CHOICE

Of the 240 patients randomized to lithium carbonate in the Bipolar CHOICE trial, a total of 145 (mean age 39.3, 59.4% women) had at least once reported the time since last lithium intake at the same visit when serum lithium levels were measured.

The 145 patients yielded 287 blood tests after 2 (N=93), 16 (N=97) and 24 (N=97) weeks. The mean time since the last lithium dose was 12.8 hours (SD=9.1; range 0.5-120) and the median time was 12 hours (IQR=10.5-14). A total of 158 (55.1%) lithium blood tests were taken between 10-14 hours after the lithium dose whereof 78 (27.2%) were taken at 12 hours, while 129 (44.9%) blood tests were taken <10 hours or >14 hours after the last lithium dose.

We found an association between a longer time since the last lithium intake and a lower lithium blood level (Figure 1A and Table 1). This was independent of the lithium dose, as we found similar lithium doses and levels of creatinine among those with <8, 8-12, 12-14, 14-16, 16-20, or >20 hours since the last lithium dose (Table 1). The creatinine concentration was higher in the three groups where blood tests were taken <14 hours since the lithium dose.

**Table 1:** Characteristics depending on the time since the last lithium dose in A) the Bipolar CHOICE trial and B) the Central Denmark Region.

|  | Time in hours since last lithium dose |  |  |  |  |  |
| --- | --- | --- | --- | --- | --- | --- |
|  | <8 (N=53) | 8-12 (N=104) | 12-14 (N=63) | 14-16 (N=24) | 16-20 (N=26) | >20 (N=17) |
| **Mean±SD lithium** | 0.64 ±0.40 | 0.67 ±0.31 | 0.59 ±0.36 | 0.45 ±0.24<sup>1</sup> | 0.44 ±0.19<sup>1</sup> | 0.29 ±0.19<sup>1</sup> |
| **Mean lithium dose, mg** | 852 ±300 | 937 ±304 | 892 ±318 | 867 ±303 | 776 ±242 | 801 ±250 |
| **Creatinine** | 0.97 ±0.22 | 0.91 ±0.21 | 0.96 ±0.28 | 0.86 ±0.14 | 0.82 ±0.11 | 0.81 ±0.18 |

**B) Central Denmark Region**
|  | Time in hours since last lithium dose |  |  |  |  |  |
| --- | --- | --- | --- | --- | --- | --- |
|  | 8-10<br>(N=1692) | 10-12<br>(N=10979) | 12-14<br>(N=15235) | 14-16<br>(N=12037) | 16-20<br>(N=8208) | >20<br>(N=3919) |
| **Mean±SD lithium** | 0.67±0.22 | 0.66±0.21 | 0.64±0.21 | 0.63±0.21 | 0.63±0.21 | 0.59±0.25 |
| **Mean lithium dose, mg** | N/A | N/A | N/A | N/A | N/A | N/A |
| **Creatinine** | 78.7±25.2<br>(n=1173) | 77.5±23.6<br>(n=7768) | 76.2±20.0<br>(n=10898) | 77.8±22.8<br>(n=8972) | 90.0±78.0<br>(n=6039) | 88.0±31.7<br>(n=2864) |
| **Mean (SD) age, years** | 48.2(16.5) | 48.9(16.7) | 47.9(17.0) | 50.1(17.2) | 52.8(17.9) | 60.2(17.4) |
| **Percentage females** | 64.1% | 66.2% | 64.7% | 61.7% | 61.4% | 62.0% |
| **Percentage private practitioners** | 27.9% | 37.8% | 36.6% | 42.1% | 45.3% | 69.0% |

Abbreviations: SD = Standard deviation.
<sup>1</sup> ANOVA with Bonferroni, Scheffe and Sidak multiple comparison corrections showed that individuals with 14-16, 16-20 and >20 hours since the last lithium dose had significantly lower lithium levels compared to individuals with 8-12 hours since the last lithium dose.

**Figure 1:**
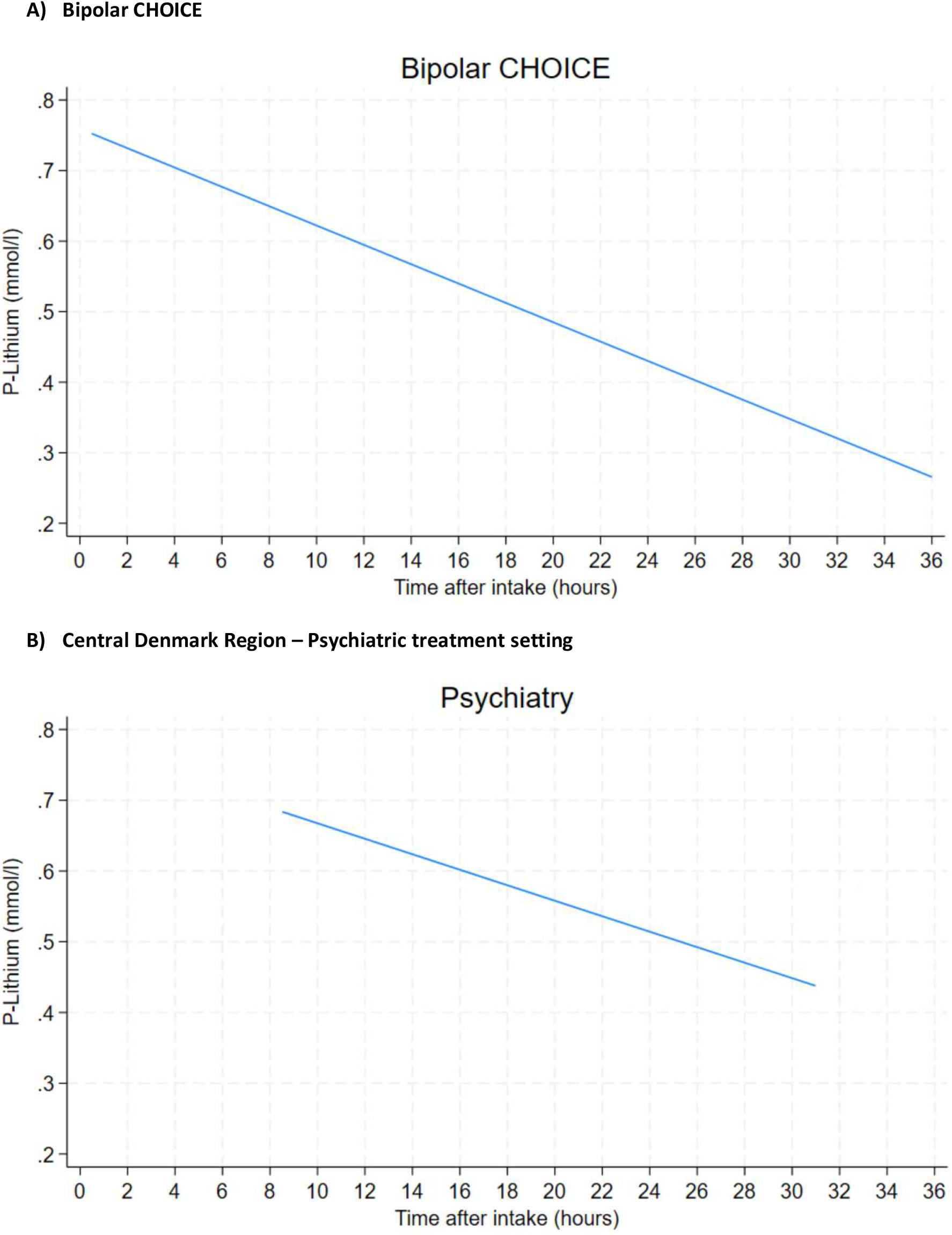
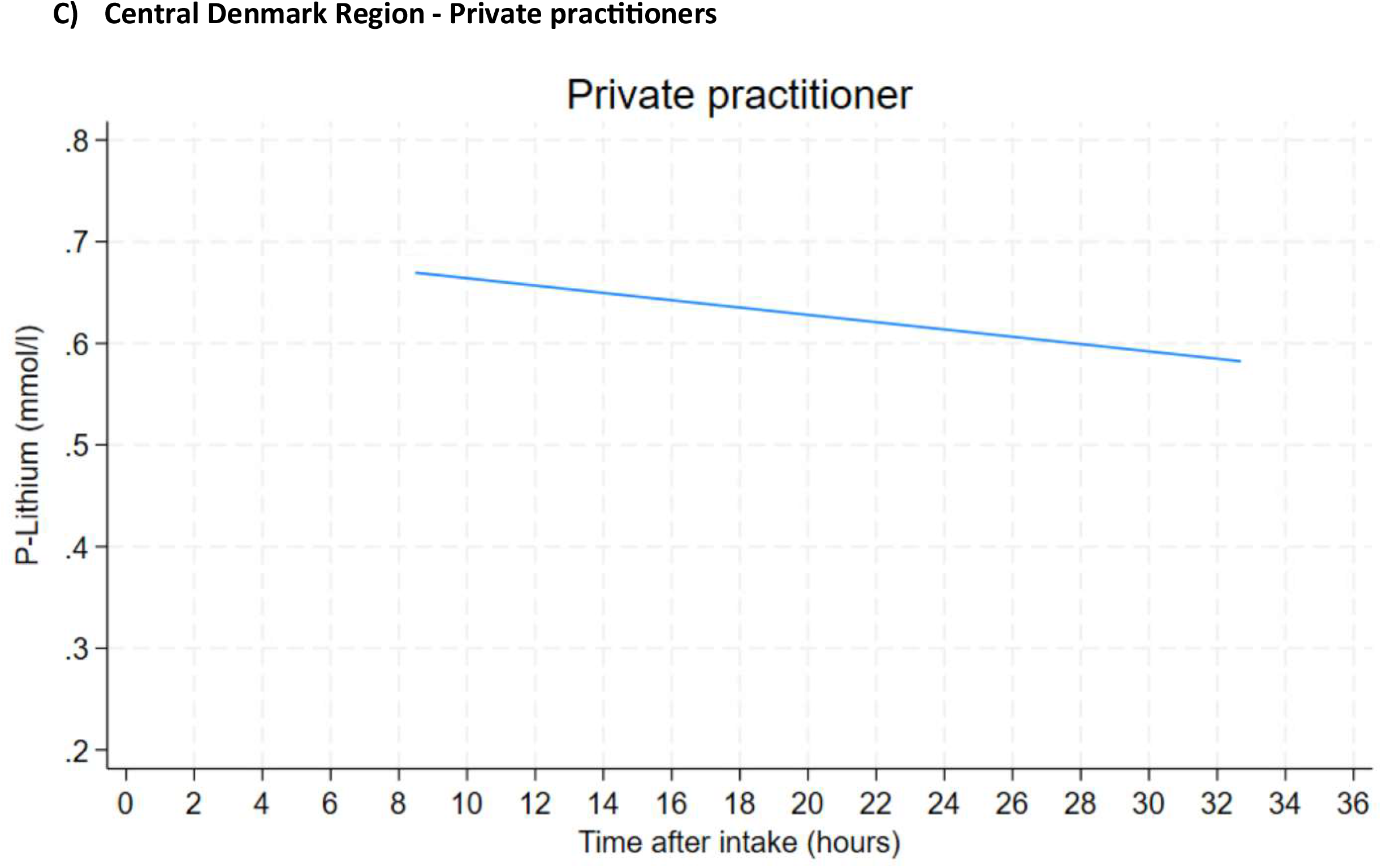
The correlation between the time since the last lithium dose and the lithium blood concentration in A) the Bipolar CHOICE clinical trial (N=291) and B) the Central Denmark Region (N=52,837), with the latter being divided into psychiatric and private practice treatment settings.

### CDR

We identified 3,177 lithium-treated individuals (mean age 45.5, 61.9% women) who had 52,070 blood tests taken from January 1, 2012, to July 31, 2022. Of the 3,177 individuals, 2,816 (88.6%) were treated with lithiumcitrate. A total of 28,432 (57.1%) lithium blood tests were taken in a psychiatric hospital setting and 21,339 (42.9%) by non-hospital settings (i.e., primarily private practitioners).

The mean time between the supposed time of lithium intake and the time stamp for the lithium blood test was 14.5 hours (SD=3.8), while the median was 13.7 hours (IQR=12.0-15.8). A total of 9,454 blood tests (18.2%) were taken approximately 12 hours after the supposed lithium intake (i.e., between 11.5 and 12.5 hours after the supposed lithium intake), while 25,856 blood tests (49.7%) were taken <10 hours or >14 hours after the last lithium dose. Of these 25,856, the vast majority (24,164, 93.5%) were taken >14 hours, as opposed to <10 hours, after the last supposed lithium intake and primarily via private practitioners. This was primarily evident when looking at blood tests taken >20 hours after lithium intake, where 69% were via private practitioners, whereas only 30% of blood tests taken 8-10 hours after intake were via private practitioners. We found no significant differences regarding age and sex (Table 1) and between calendar years (sTable 1).

The mean lithium blood concentration decreased with longer time since the last supposed lithium intake (Figure 1B). This was more pronounced for lithium blood tests ordered in a specialized psychiatric setting (e.g., mean 12-hour level of 0.66 and mean 20-hour level of 0.56), while among blood tests ordered by private practitioners, the concentrations were almost identical over time (e.g., mean 12-hour level 0.66 and mean 20-hour level 0.63). Regarding creatinine, we found an increase depending on a longer time between the supposed lithium intake and the lithium blood test (Table 1B). Those with 16-20 hours between lithium intake and the blood test had a mean creatinine of 90, while those <14 hours had a mean creatinine between 76 and 78. The mean age was similar between these groups.

Next, we illustrated the time between the supposed lithium intake and the lithium blood test depending on the supposed time point for the lithium intake (Figure 2). We found a clear pattern that when the supposed lithium intake was earlier than 8 pm, most lithium blood tests were taken >14 hours after the last supposed lithium intake. Patients who were supposed to take their lithium medication at 9, 10 or 11 pm, had a mean of approximately 12 hours between the lithium intake and the lithium blood test.

**Figure 2:**
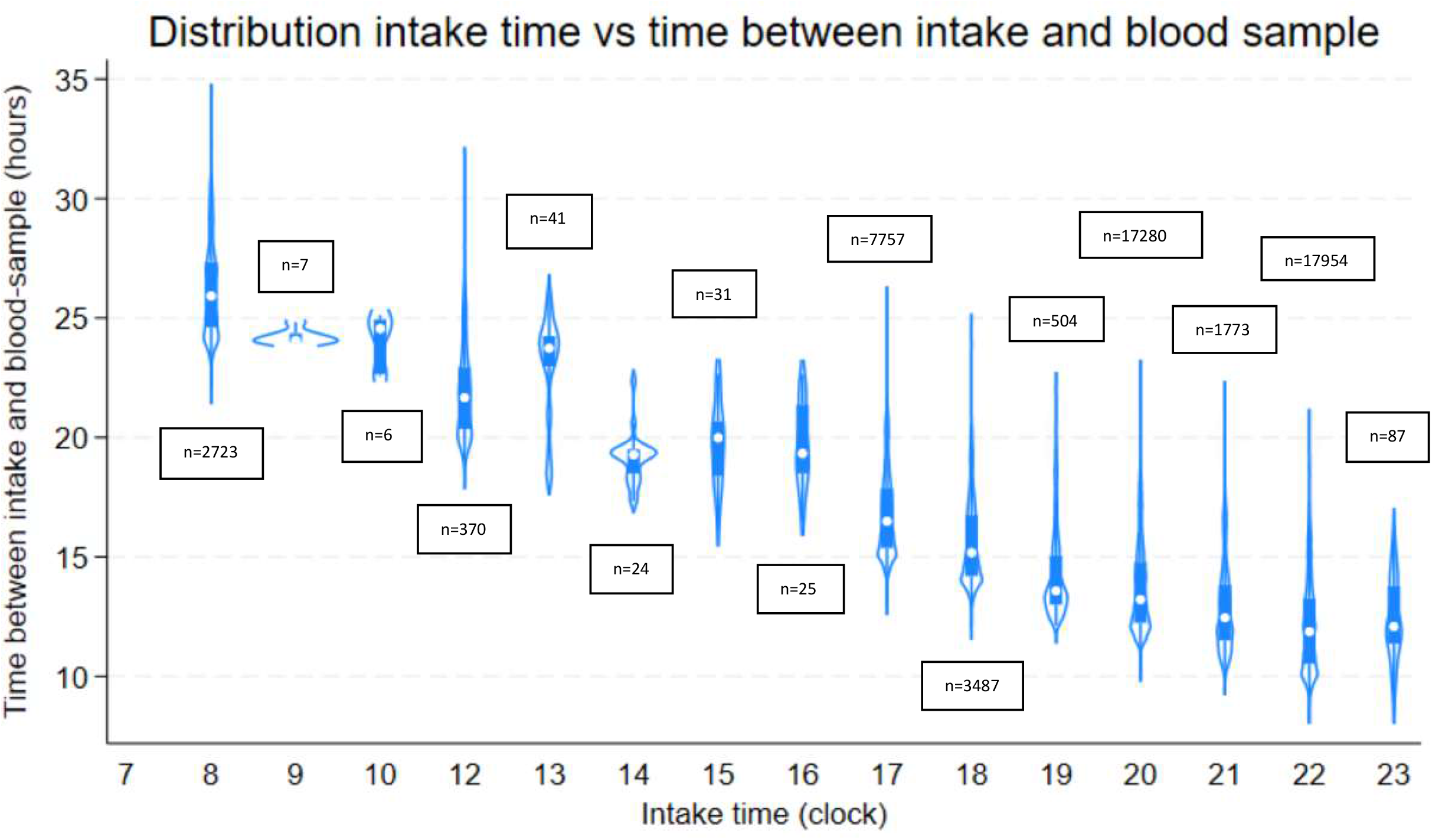
Among 3,177 patients in the CDR, showing the correlation between the time when patients were supposed to take their lithium medication (x-axis) and the time in hours between the lithium intake and the registered time stamp of the lithium blood test (y-axis).

## Discussion

This study has three main findings. First, approximately half of lithium blood tests substantially deviate from the recommended 12-hour post-dose timing, posing a clear risk of negatively impacting clinical decision-making. Second, our data indicate that lithium-treated patients under the care of general practitioners (who made up the vast majority of private practitioners in this study), to a lesser degree comply with the 12-hour timing. Third, the latter seems to be associated with higher creatinine levels, which could indicate that lithium-treated patients under the care of general practitioners are at higher risk of long-term kidney damage due to higher lithium concentrations.

Hence, the most important finding from this study is that many lithium blood tests do not comply with the 12-hour timing. We found very similar numbers in the clinical trial with self-reported data and the observational study with estimations based on registry data (i.e., 45% and 50%), indicating that approximately half of lithium blood tests are drawn outside the recommended interval of 10-14 hours after the lithium dose. Only a minority (27% and 18%, respectively) of lithium blood tests were taken 12 hours after the last lithium dose. This has clear clinical implications, as the 12-hour lithium blood level represents an important tool to the clinician when determining a patients lithium dose. As also shown by the present data, a deviation from the 12-hour timing can result in rather large deviations in lithium blood concentrations. This is particularly important as many patients continue with lithium treatment for many years or even decades. In Denmark, lithium treatment should only be initiated by psychiatrists, but patients are usually treated in specialized psychiatric settings (e.g., a bipolar disorder outpatient clinic) for less than two years. After this, patients are mostly followed by their general practitioners (in Denmark, there are few private practicing psychiatrists), who have less expertise in lithium treatment. Hence, there is a risk that general practitioners are not aware of the necessity of the 12-hour blood test timing, potentially resulting in lithium dosages based on, and not adjusted for, incorrectly timed blood tests.

We found interesting differences in creatinine concentrations between the clinical and the observational datasets. In Bipolar CHOICE, we found slightly higher creatinine levels among blood tests taken <14 hours after the last lithium dose, with these blood tests also showing higher lithium concentrations. This seems reasonable, as higher lithium concentrations directly affect kidney functioning and hence explain the higher creatinine levels. However, in the observational dataset, we found the opposite trend. Those with >16 hours between lithium intake and the lithium blood test had higher creatinine levels. In addition, we did not observe a similar decrease in lithium concentration among those blood tests taken >16 hours after the last lithium dose. A subgrouping into specialized hospital settings versus non-hospital settings showed that blood tests taken >16 hours after the last lithium dose were particularly often ordered by private practitioners. This could indicate that lithium is dosed based on a falsely low lithium blood concentration by clinicians with less expertise in lithium treatment (e.g., general practitioners interpreting an 18-hour level as the 12-hour level and prescribing higher lithium blood concentrations based on this falsely low lithium concentration), leading to higher creatinine levels over time. As lithium treatment often lasts for several years or even decades, this may result in an increased risk for long-term kidney damage.

### Strengths and limitations

The primary strength of this study is the use of data from a controlled clinical trial and a large observational setting representing real-world clinical practice, showing similar numbers in settings with self-reported time between lithium intake and the blood test and when estimating this based on registered time stamps.

The primary limitation of the CDR data is that we have no information on the time for the actual intake of the lithium medication, but only the registered time point for when the patient is supposed to take the medication. The fact that the lithium blood concentration only decreased slightly with the time since last lithium dose in the CDR data may be explained by that the registered time does not represent the actual time for lithium intake and/or missing knowledge by clinicians and patients on the necessity for 12-hour timing. Indeed, as the registered time for lithium intake may not represent the actual time for lithium intake (e.g., registered for intake at 6 pm but the actual intake by the patient is at 8 pm), a blood test coded as being 17 hours after the last lithium dose may actually represent a blood test that is closer to the 12-hour level. Therefore, we do not know whether patients actually took their lithium at the registered time or at other time points that fit better with 12-hour timing. Nevertheless, it is reassuring that the numbers are highly similar to the clinical trial with self-reported data.

## Conclusion

Approximately half of lithium blood tests do not comply with the 12-hour timing, although the 12-hour trough level has been the gold standard for informing clinicians about individual lithium blood concentrations for >50 years. This increases the risk for unprecise clinical decisions on lithium dosage that could impact individual patients, e.g., prescribing a higher lithium dose based on a presumed 12-hour lithium blood concentration that is falsely low. Furthermore, the present data indicate that non-compliance with 12-hour timing seems to be particularly frequent among general practitioners and may correlate with higher creatinine levels, which may affect long-term risks on the kidneys. These findings, therefore, emphasize the need to improve the clinical management of lithium blood level testing, which would have the potential to optimize the long-term treatment and follow-up of lithium-treated patients.

## Data Availability

All data produced in the present study are available upon reasonable request to the authors

---

**Supplementary Table 1:** Presenting the time between the last supposed lithium intake and the lithium blood test depending on calendar year of the lithium blood test.

|  | 2012 | 2013 | 2014 | 2015 | 2016 | 2017 | 2018 | 2019 | 2020 | 2021 | 2022* |
| --- | --- | --- | --- | --- | --- | --- | --- | --- | --- | --- | --- |
| **NUMBER OF SAMPLES** | 1612 | 2038 | 2997 | 3787 | 5056 | 5673 | 6006 | 6765 | 7144 | 7192 | 3800 |
| MEAN TIME (SD) BETWEEN LAST SUPPOSED LITHIUM INTAKE AND LITHIUM BLOOD TEST | 14.51<br>(0.09) | 14.61<br>(0.08) | 14.62<br>(0.07) | 14.55<br>(0.06) | 14.75<br>(0.05) | 14.53<br>(0.05) | 14.58<br>(0.05) | 14.35<br>(0.04) | 14.26<br>(0.04) | 14.40<br>(0.04) | 14.32<br>(0.06) |
| PERCENTAGE OF LITHIUM BLOOD TESTS TAKEN <10 OR >14 HOURS AFTER THE LAST SUPPOSED LITHIUM INTAKE | 52.11<br>% | 50.10<br>% | 48.62<br>% | 50.49<br>% | 47.55<br>% | 47.61<br>% | 48.82<br>% | 51.34<br>% | 52.53<br>% | 51.35<br>% | 53.47<br>% |

Abbreviations: SD = Standard deviation.
\* In 2022, data were available until July 31.

## Notes

### Competing Interest Statement

The authors have declared no competing interest.

### Funding Statement

This study was funded by an unlimited grant by the LundbeckFoundation

### Author Declarations

Approval for the present study was granted by the Legal Office in the Central Denmark Region in accordance with the Danish Health Care Act (Section Sign)46, Section 2. According to Danish Law, patient consent and approval from ethical/institutional review boards are not required for studies of this type. Bipolar CHOICE Trial: Subjects provided verbal and written informed consent prior to participation and all Institutional Review Boards of the different sites approved the study protocol.

